# Interprofessional Care Team, Staffing, and Setting Characteristics that Impact Patient Outcomes: A Review of Reviews

**DOI:** 10.1101/2024.01.04.24300868

**Authors:** Alix Pletcher, Kyla Woodward, Natalie Hoge, Nathaniel Blair-Stahn, Paulina Lindstedt, Zahra Gohari, Abraham Flaxman, Sarah Iribarren

## Abstract

**Background:** The purpose of this study was to identify research methods and evidence pertaining to the relationship of interprofessional acute care teams and hospital characteristics on patient outcomes in hospital-based acute care.

**Methods:** A review was completed using the Preferred Reporting Items for Systematic Reviews and Meta-Analysis extension for Scoping Reviews guidelines. The search strategy was executed across PubMed, CINAHL, and Embase. The review included 12 systematic reviews from 2012 to 2023 that examine the impact of acute care staffing characteristics on patient outcomes.

**Results:** Workforce characteristics primarily focused on nurse staffing, with a limited number of studies assessing the impact of interprofessional teams or non-clinical workers on care quality. There is limited data describing the context of care delivery via potential relationships between hospital characteristics, interprofessional team staffing levels, and patient outcomes.

**Conclusions:** To promote comparability across studies, future workforce research should include a comprehensive analytic approach that includes clearly defined variables representing interprofessional care teams, community factors, and staffing and patient characteristics.

## Introduction and Background

Shortages of healthcare workers have been a long-standing and pervasive issue in international acute care settings, but since the onset of the COVID-19 pandemic, maintaining adequate staffing levels has become an even greater challenge for acute care facilities. Internationally, healthcare workers left jobs or reduced hours for multiple reasons, including illness, fear of infecting themselves or loved ones with COVID-19, needing to care for children and other family members, and the effects of extended heavy workloads.(1–3) Those who stayed at work also experienced persistent negative impacts, with one international review identifying consequences such as losing hope or professional identity,(4) which have led to persistent workforce shortages.)

Ongoing staffing shortages place a burden on all healthcare workers and affect their perceptions of safety in the workplace,(5) and also impact patient, particularly those with higher support needs related to individual. A number of studies have demonstrated that nurse staffing and adverse patient outcomes are inversely related, so that higher staffing ratios result in fewer adverse outcomes.(6, 7) However, despite the fact that nursing care does not occur in a void, there is little if any data exploring links between nursing workload, nurse burnout, and staffing of other interprofessional care team members. Nurse staffing data does not provide information about the staffing of other clinical and nonclinical care team members such as social workers, physical and respiratory therapists, nutrition services, and environmental services, collectively called the ‘interprofessional care team’, who help provide critical support and services for patients with higher needs. While workers in these roles may not provide direct care, their jobs include tasks often absorbed by nursing staff when there is insufficient staffing. This absorption of additional duties contributes to burnout and dilution of RN scope of practice, increasing the likelihood of missed care and other adverse events, and leading to missed opportunities to advance equity.(8, 9)

Likewise, few studies account for specific features of communities and patients that impact acute care experiences and outcomes, particularly regarding health equity. Community level factors such as urbanicity, county socioeconomic status, and housing type may shape the patient population served by the organization and the resources available to patients after discharge. Patient outcomes also differ due to characteristics such as comorbidities, mental health problems, and social determinants of health (SDOH), a set of social and environmental factors underlying health inequities.(10) Increases in inpatient assessment of SDOH have improved the care teams’ ability to identify patients with higher social and support needs; those needs require interprofessional services and support alongside nursing care to improve equity and optimize patient outcomes.(11) With ongoing shortages of healthcare workers across multiple professions and documented burnout among RNs, it is critical to understand how care team composition in acute care settings affects outcomes for both patients and workers.

In 2021, the WA legislature directed the Washington State Department of Health to contract an interdisciplinary team of University of Washington researchers led by the School of Nursing to conduct a workforce study examining the impacts of healthcare staffing characteristics on patient outcomes.(12) As our first step in developing an appropriate analytic strategy for the study, this paper reports a review of reviews of existing international data on factors that may influence both care team staffing and patient outcomes. The purpose of this study was to identify what variables have been used to examine relationships between interprofessional team staffing and patient outcomes, including variables pertaining to community, hospital, care team, and patient characteristics that may influence either staffing or outcomes, and to develop a preliminary causal model for further use in our project.

## Methods

This study focused on identifying variables used to assess the impact of inpatient care teams on patient outcomes. We mapped the types of variables, outcomes, and analyses used to evaluate relationships between interprofessional acute care teams and patient outcomes in existing literature. We chose to review systematic reviews to allow representation of a large pool of literature and get a high-level overview of what has been studied in this domain to ascertain the scope of the research and identify gaps in knowledge.(13) The stages of the review included establishing research question(s), identifying relevant studies, selecting studies, charting the data, and collating, summarizing, and reporting the results.(14) This study was designed and reported according to the Preferred Reporting Items for Systematic Reviews and Meta-Analysis (PRISMA) Guidelines.(15)

### Step 1. Establish the research questions

Using our team’s expertise and directives from the legislature, we narrowed our research questions to the following:

1. How were team members from different professions and roles (clinical and nonclinical) represented in studies assessing the impact of care teams on patient outcomes?
2. What community, hospital/organizational, care team, and patient factors were included in description or analysis of staffing as it impacted patient outcomes, and how were these variables defined?
3. What analytic strategies were used to examine the impact of teams or staffing on patient outcomes?

### Step 2. Identify relevant studies

The search strategy was developed and executed in consultation with an experienced research librarian (CM). The original search occurred on March 3, 2022 and was most recently updated on March 30, 2023. The search strategy was created for PubMed (which includes Medline, PubMed Central, and other resources) using a combination of Medical Subject Heading (MeSH) terms, keywords, and phrases (see the complete search strategy in Supporting Information). The search terms targeted health workers, health research, hospitals/hospital settings/hospitalization, and patient outcomes. The PubMed search terms were translated for Cumulated Index to Nursing and Allied Health Literature (CINAHL) and Embase using their respective thesaurus terms and advanced search features. A manual search using the reference lists of retrieved citations was conducted for other relevant studies.

Table 1 summarizes the participants, interventions, comparison, outcomes, and study design (PICOS) used to define the inclusion and exclusion criteria strategy. Eligible studies included systematic reviews that examined interventions with a workforce component and their impact on adult acute care patient outcomes and were published within the last 10 years. We excluded all other study or publication types, such as cross-sectional studies, randomized control trials, commentaries, editorials, letters to the editor, abstract proceedings and reviews that did not include workforce personnel or patient outcomes. We restricted our search to only include reports that were published in English.

**Table 1.** Study inclusion criteria.

| PICOS element | Description |
| --- | --- |
| Population | Adults in acute care hospitals |
| Intervention or Comparison | Staffing models or measures, including those focused on nurses or other interprofessional care team members OR Staff training or education level |
| Outcome | Any patient outcome (e.g., mortality, pressure ulcers) |
| Study type | Systematic review, with or without meta-analysis |

### 3. Select Relevant Studies

We compiled the search results in EndNote and removed duplicates, then transferred citations to Covidence software for screening. The screening process was composed of two phases: first, we reviewed the title and abstract for relevance using the inclusion criteria; and second, we reviewed the full text of remaining articles. In each of these phases, two reviewers (AP, ZG) independently screened the articles and a third reviewer (SI) assisted in resolving any discrepancies.

### 4. Charting the Data

Project directives informed the selection of variables to include analysis of the impact of the number, type, education, training, and experience of acute care hospital staffing personnel on patient mortality and patient outcomes. Control variables included factors such as access to equipment, patients’ underlying conditions and diagnoses, patients’ demographic information, the trauma level designation of the hospital, transfers from other hospitals, and external factors impacting hospital volumes.(12) Based on an initial review of papers to identify key components, the reviewers used a standard data extraction sheet to record the following categories: study design, data sources, inclusion criteria within that review, interprofessional team members, types of patients (e.g. acute versus intensive care), patient outcomes (e.g., length of stay, hospital acquired infection), additional factors considered in the review or analysis (e.g., equipment, technology, external factors), and analytic strategy or model. In addition, any reported challenges, limitations, strengths, or recommendations to each were extracted.

### 5. Synthesis of Results

We used a descriptive approach to report the data on the variables, approaches, and healthcare team composition. Given the expected heterogeneity of systematic reviews found in our search and our goal to simply identify the presence and use of variables rather than the direction of any relationships between them, a quantitative summary measure of results was not undertaken. We summarized frequency of use, definitions, and concordance of analytical strategies across the studies. We synthesized the findings into main concepts, grouped by variable categories (i.e., hospital characteristics, patient characteristics, staffing, or patient outcome). We also summarized reported challenges, limitations, strengths, and recommendations for each variable. Results informed the development of a baseline causal model that was used to guide subsequent stakeholder interviews and to develop and refine the analytic strategy for the state workforce study.

## Results

Search results yielded a total of 164 articles, of which 37 met the inclusion criteria for full text review (see Figure 1). We then identified a total of 12 systematic reviews for data extraction, which represented more than 575 individual studies (not counting duplicate instances of 13 studies among the reviews). Only 4 reviews included data from the most recent 5 years,(16–19) while the remainder included studies dating back to 1986. One review focused only on the United States,(8) and the other eleven included international data. Reviews predominantly included reports of primary research, with only one using secondary sources such as reviews or editorials.(19) We organized results into interprofessional team and staffing characteristics, elements of communities, hospitals and patients that may interact with staffing characteristics, and patient outcomes (see Table 2). We highlighted elements that consider personnel or factors outside of nursing to understand the relationship between healthcare teams and patient outcomes.

**Figure 1.**
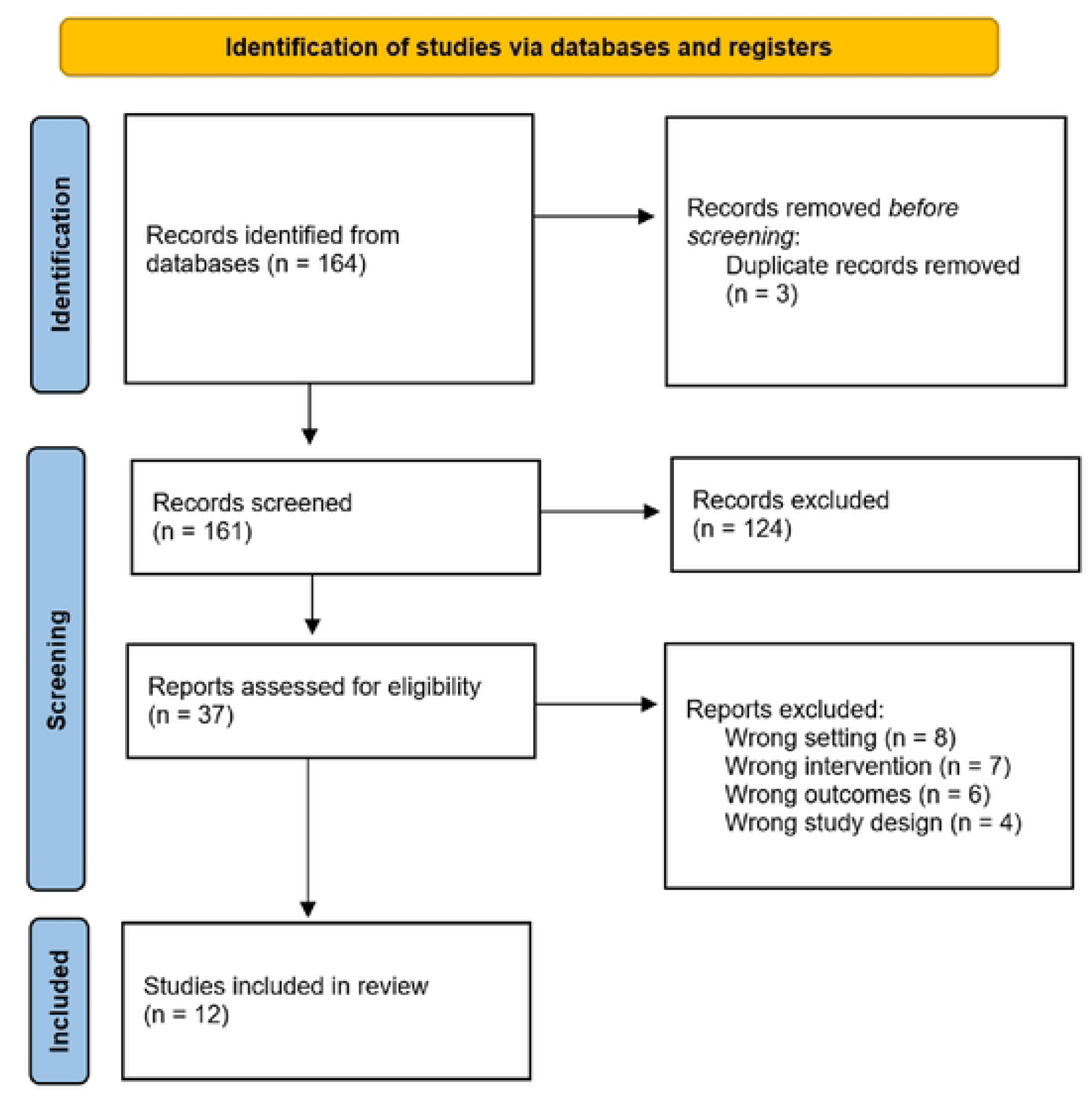
PRISMA flow diagram.

**Table 2.** Overview of included studies.

| Review<br>(# studies) | Study aim | Staffing variables | Patient outcome<br>variables | Other variables |
| --- | --- | --- | --- | --- |
| **Bae et al.**<br>**(2014)(26)**<br>**n=24** | To systematically evaluate the effect of nurse overtime and long work hours on nurse and patient outcomes | RN work hours<br>RN overtime (OT) | Adverse events<br>Failure-to-rescue<br>Mortality<br>Patient satisfaction | N/A |
| **Bagnasco et al.**<br>**(2019) (16)**<br>**n=44** | To review and synthesize research studies on surgical and medical inpatients' perceptions on unmet nursing care needs | Nursing workload<br>NHPPD<br>Nursing skill mix<br>RN OT | Missed patient care<br>Mortality | Team: work environment, RN education, RN experience<br>Hospital: size, technology level, teaching status<br>Patient: age, sex, admission type, comorbidities |
| **Burke et al. (2021)**<br>**(23)**<br>**n=52** | To investigate what clinically relevant interventions have been shown to improve organizational fail-to-rescue rates | NPR | Failure-to-rescue<br>Mortality | Team: work environment; RN education |
| **Cassarino et al.**<br>**(2019)(20)**<br>**n=6** | To synthesize the totality of evidence relating to the impact of early assessment and intervention by healthcare teams on quality, safety, and effectiveness of care in the Emergency Department | Presence of interdisciplinary team members | Hospital admission<br>Length of stay<br>Mortality<br>Patient satisfaction<br>Readmissions | N/A |
| **Dall'Ora et al.**<br>**(2022)(17)**<br>**n=27** | To evaluate evidence for an association between nurse staffing levels and patient outcomes in acute care settings | NPR<br>NHPPD | Adverse events<br>Length of stay<br>Mortality<br>Readmissions | Team: RN skill mix<br>Patient: age, sex, admission type, comorbidities, severity of illness |
| **Evangelou et al.**<br>**(2018)(22)** | To identify quality indicators associated with | NPR; NHPPD<br>Nursing manpower use score | Adverse events<br>Cost<br>Length of stay | Team: work environment, RN education, RN |
| **n=13** | nursing care for adult ICUs in literature | Patient care assistant-to-patient ratio | Mortality<br>Patient satisfaction<br>Readmissions | experience, RN skill mix, RN group competence |
| **Griffiths et al. (2018)(8)**<br>**n=18** | To identify nursing care most frequently missed in acute adult inpatient care wards and to determine evidence for the association of missed care with nurse staffing | NPR | Missed patient care | N/A |
| **Kerlin et al. (2016)(21)**<br>**n=18** | To review the association of nighttime intensivist staffing with outcomes of ICU patients | Nighttime intensivist staffing<br>Nighttime:daytime staffing differences<br>Staff type | Complications<br>Duration of mechanical ventilation<br>Length of stay<br>Mortality<br>Readmissions | Hospital: size<br>Patient: severity of illness |
| **McGahan et al. (2012)(24)**<br>**n=19** | To examine the relationship between nurse staffing levels and the incidence of mortality and morbidity in adult intensive care unit patients | NPR; NHPPD<br>Therapeutic Intervention Scoring System-to-RN ratio<br>Bed-to-nurse ratio | Adverse events<br>Mortality | N/A |
| **Plotnikoff et al. (2021)(19)**<br>**n=314** | To investigate what elements facilitate a successful, high-quality discharge from the ICU | NPR<br>Provider experience; provider training; provider workload; presence of an interdisciplinary team | Adverse events<br>Costs<br>Length of stay<br>Mortality<br>Patient satisfaction<br>Readmissions | Team: provider-to-provider communication<br>Patient: demographics, provider-to-patient communication, family engagement, discharge education; admission type<br>Hospital: trauma designation, technology |
| **Rae et al. (2021)(18)**<br>**n=55** | To determine associations between variations in registered nurse staffing levels in adult critical care | NPR; NHPPD<br>Bed-to-nurse ratio<br>Number of RNs<br>Nursing Activities Score | Adverse events<br>Mortality<br>Length of stay<br>Duration of mechanical ventilation | N/A |
|  | units and outcomes such as patient, nurse, organizational and family outcomes | RNs' perception of staffing adequacy | Length of weaning<br>Patient satisfaction |  |
| **Recio-Saucedo et al. (2017)(25)**<br>**n=14** | To systematically review the impact of missed nursing care on patient outcomes, in acute hospital wards and nursing homes | NPR; NHPPD | Adverse events<br>Length of stay<br>Mortality<br>Patient satisfaction<br>Patient safety<br>Readmissions | Team: work environment, RN education, RN experience, RN age<br>Hospital: size, teaching status, Magnet© designation, type, technology level, ownership, Medicare cost-to-charge ratio<br>Community: population density, volume of patients with heart failure, hospital location, language region, profit status<br>Patients: demographics, admission type, insurance status, number of procedures, patient health status, comorbidities |

Note. RN = Registered Nurse; ICU = Intensive Care Unit; NPR = Nurse-to-Patient Ratio; NHPPD = Nursing Hours Per Patient Day

**Table 3.**
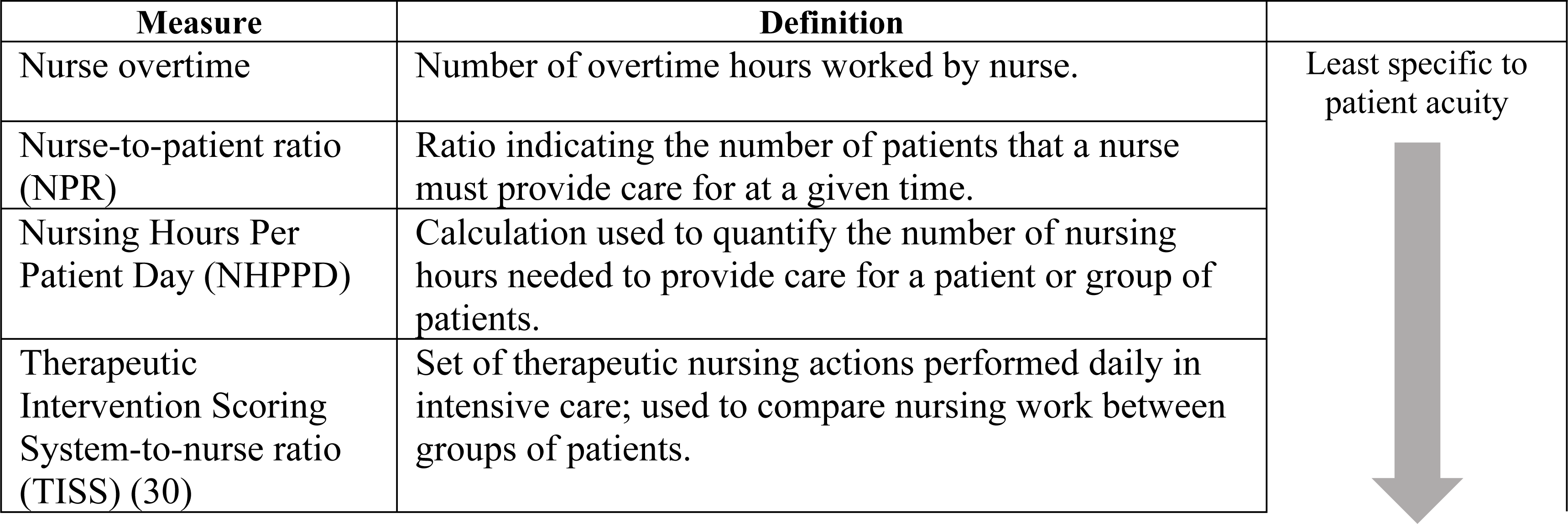
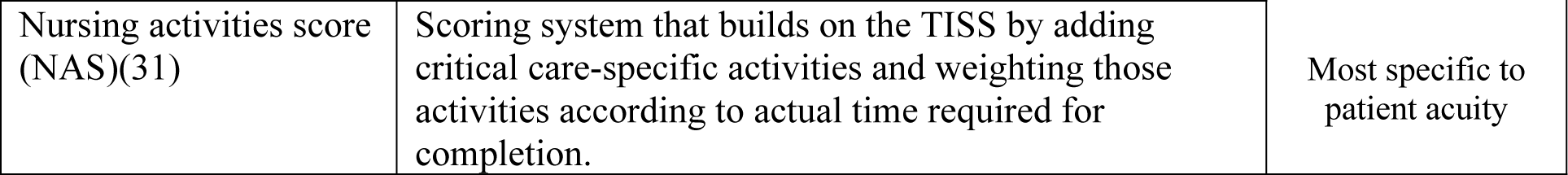
Nursing workforce measures and definitions.

| Measure | Definition |  |
| --- | --- | --- |
| Nurse overtime | Number of overtime hours worked by nurse. | Least specific to patient acuity |
| Nurse-to-patient ratio (NPR) | Ratio indicating the number of patients that a nurse must provide care for at a given time. |  |
| Nursing Hours Per Patient Day (NHPPD) | Calculation used to quantify the number of nursing hours needed to provide care for a patient or group of patients. |  |
| Therapeutic Intervention Scoring System-to-nurse ratio (TISS) (30) | Set of therapeutic nursing actions performed daily in intensive care; used to compare nursing work between groups of patients. |  |
| Nursing activities score (NAS)(31) | Scoring system that builds on the TISS by adding critical care-specific activities and weighting those activities according to actual time required for completion. | Most specific to patient acuity |

### Interprofessional Teams and Staffing

The reviews provide limited evidence on the impact of interprofessional healthcare teams on patient outcomes. Most studies either focused exclusively on nursing staff (10/12) or considered direct care providers such as physicians or respiratory therapists (2/12) without mentioning the impact of indirect healthcare workers such as nutrition or environmental services. One study of interprofessional teams in the emergency department defined the healthcare team as an assortment of care professionals including occupational and physical therapists and medical social workers.(20) One review examined the relationship between nighttime intensivist staffing and patient outcomes in intensive care.(21) Two reviews included a limited number of studies examining relationships between non-registered nurse team members and quality indicators or discharge.(19, 22)

Studies that quantified the impact of nursing staff on patient outcomes used various measures to quantify the patient care workload. The most common measure of absolute staffing levels was nurse-to-patient ratio (8/12),(8, 17–19, 22–25) followed by Nursing Hours Per Patient Day (6/12).(16–18, 22, 24, 25) Other less commonly used measures for nursing workload included nurse overtime (2/12),(16, 26) Nursing Activities Score (1/12),(18) number of hours worked by nurses (1/12),(26) and Therapeutic Intervention Scoring System-to-nurse ratio (1/12).(24)

### Nurse characteristics

As part of assessing staffing, reviews addressed characteristics of nursing staff and workplace factors. Assessments of nursing work environment (as measured by the Nursing Work Scale) were included in 4/12 reviews.(16, 22, 23, 25) Another common characteristic was education (4/12), or the percentage of RNs with a baccalaureate or higher degree in nursing.(16, 22, 23, 25) Three of those reviews also looked at years of nursing experience,(16, 22, 25) 2/12 included specialty certification, (22, 25) and 1/12 included tenure at the organization.(25) Job-related training or knowledge was included in 3/12 reviews,(17, 19, 25) 3/12 looked at nursing skill mix, or the percentage of nursing staff who were registered nurses,(16, 17, 22) and 1/12 included nurse reported quality measures.(25) Most studies used these elements as independent variables, while 1/12 used them as controls for their main findings.(19)

### Community, Hospital, and Patient Characteristics

In general, community characteristics such as rurality or population demographics were not used as independent variables or controls when relationships between healthcare teams and patient outcomes were assessed; only 1/12 reviews included community characteristics as external factors that may affect hospital volume.(25) Within hospitals, characteristics such as equipment and technology, size, and teaching status were examined in 4/12 reviews.(16, 17, 19, 25) Two reviews controlled for equipment and technology, with definitions ranging from counts of resources such as critical care beds to facilities for major organ transplant or open-heart surgery.(16, 25) One review included hospital-level structures for discharge.(19) In all, community and hospital variables were not consistently represented or defined across the included reviews.

Patient-level characteristics were used as descriptive and control variables. 4/12 reviews considered measures related to patient health status at baseline or severity of illness during hospitalization.(17, 19, 21, 25) Only 2/12 reviews addressed other patient variables such as demographics and healthcare logistics such as insurance or finances(19, 25) and 1/12 looked at admission type (e.g., elective versus emergent admissions).(17) No included studies referenced or defined SDOH or discussed health equity.

### Patient Outcomes

Patient outcomes were treated as a dependent variable in most of the included reviews (9/12), and were defined as mortality, adverse events, and other care outcomes. We used the National Quality Forum classification for adverse events, which defines 29 possible serious, reportable events that are considered largely preventable and therefore indicative of inadequate healthcare safety mechanisms.(27) Patient mortality was included in some form in 11/12 reviews. Of those, 5/12 looked at inpatient mortality,(17, 18, 21, 22, 25) 5/12 looked at post-discharge mortality at 1-month(17, 18, 20, 23, 26) or 1/12 at 1 year,(20) 1/12 did not specify a timeframe,(24) and 1/12 noted the use of mortality in many of the included studies but did not treat it as a predictor or outcome.(19) Non-fatal adverse events related to inpatient care were used in 7/12 reviews as a staffing-related outcome. Reported adverse events included healthcare associated infection (6/12),(17, 18, 22, 24–26) pressure injury (5/12),(17, 22, 24–26) patient falls (3/12),(22, 25, 26) medication errors (2/12),(22, 25) and gastrointestinal bleeds (1/12).(26) Other patient outcomes were length of stay (7/12),(17–22, 25) hospital readmission (6/12), (17, 19–22, 25) satisfaction (5/12), (18, 20, 22, 25, 26) and ventilator days (3/12).(17, 18, 21) Two studies focused on missed care(8, 16) and another included health-related quality of life as a patient outcome.(20)

### Strategies for Analysis

Quantitative strategies for analysis were only addressed in 5/12 reviews. Of the reviews that extracted information on quantitative analysis, multivariate logistic regression was the most used model type in the evaluation of staffing impacts on patient outcomes (3/12),(17, 23, 26) followed by multivariate linear regression (2/12)(19, 24) and negative binomial regression (2/12).(17, 26) One of 12 reviews reported use of other statistical analyses, including the Cox proportional hazard model and hierarchical mixed effects survival.(17)

### Causal Model

Study findings were used to develop an initial causal model that was used throughout the WA workforce study the Washington Acute Care CHaracteristics and patient Outcomes (WACCHO) model (Figure 2). The model presents the different categories of variables present in the current literature. We used this model to guide our stakeholder interviews and focus groups and determine if any factors were missing or inadequately represented in the model.

**Figure 2.**
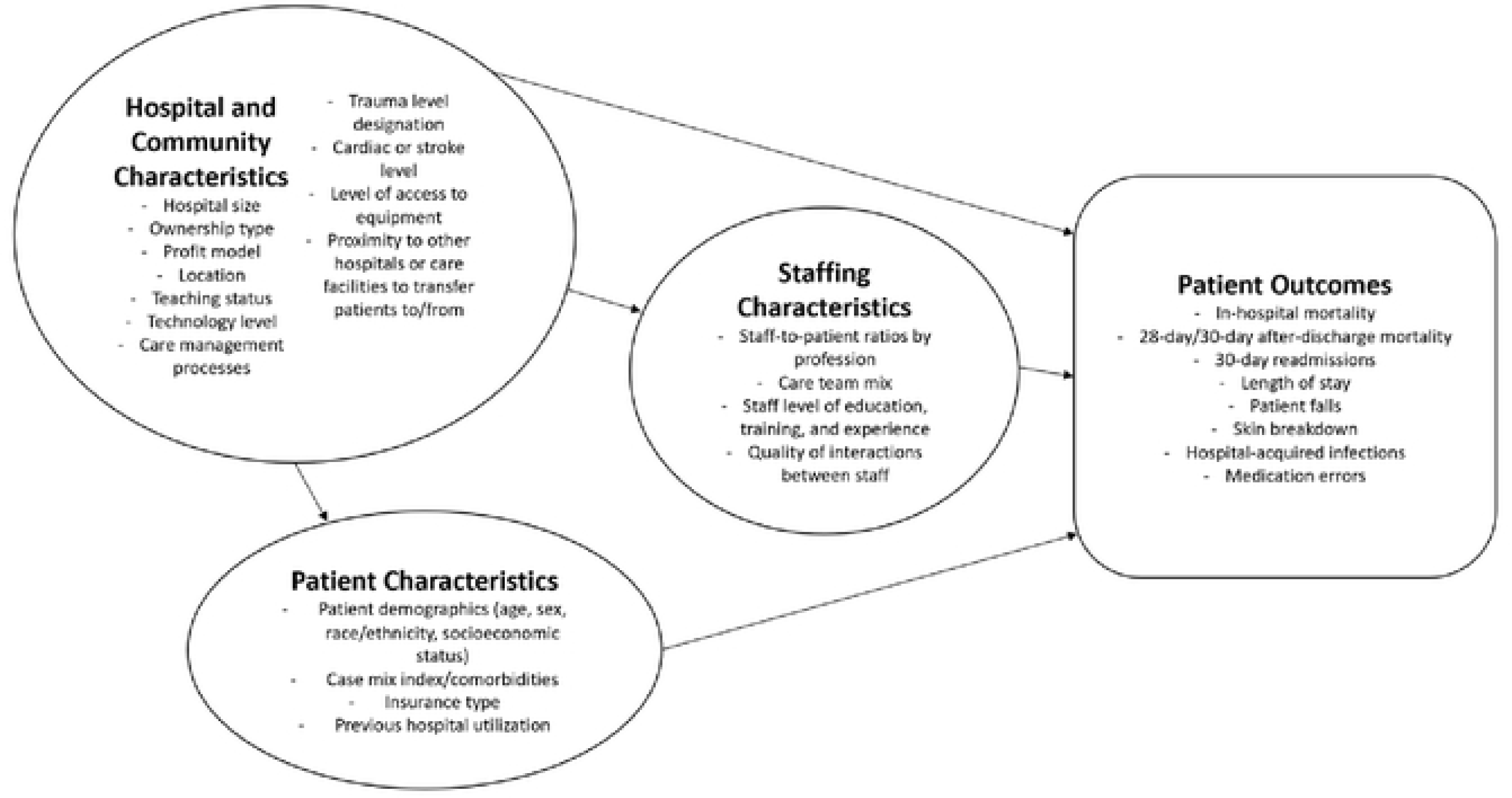
Initial causal model based on review findings.

## Discussion

Key findings in this review reinforce the reliance on nursing in existing research assessing the impact of staffing on patient outcomes. Our results also indicate a lack of attention to the contexts in which acute care occurs within organizations and the community, and which may impact patient needs and equitable care delivery. These findings suggest several critical areas for future research on staffing and patient outcomes, including interprofessional care team composition and characteristics of communities, hospitals, and patients that may influence staffing and/or patient outcomes. In particular, a lack of attention to patients’ SDOH limits our ability to understand implications of staffing on health equity. Findings also point to a need for clearly defined, consistently used variables and appropriate analytic strategies in healthcare workforce research.

### Care team composition

A key finding from this review was that interprofessional acute care team composition is undertheorized and underrepresented in current scientific literature. Because studies that included interprofessional team members occurred in specific areas outside of acute care or focused on a specific event such as discharge, we found no evidence describing the effects of a comprehensive, interprofessional care team on patient outcomes in acute care. Furthermore, studies predominantly focus on the effects of nurse staffing and infrequently consider the influence of non-nursing staff or indirect care providers such as environmental services on patient outcomes. These findings reinforce trends in current workforce analysis to focus exclusively on nursing personnel without acknowledgement of associated factors that impact the RN workload.(7) While a focus on nursing is warranted due to their essential role in acute care, research that fails to account for other factors and team members impacting RN work will miss important elements that influence the relationship between staffing and patient outcomes. For example, in the absence of other care team members, RNs often take on non-nursing roles, diluting time that could be spent on activities that optimize their full scope of practice and increasing frustration and burnout.(9) Understanding the impacts of other types of staffing is therefore essential to understanding how outcomes are affected in settings where RNs are able to exclusively focus on RN work and settings where they routinely take on other tasks. A national workplace-focused survey in the United States found that among 5,461 acute care RN respondents, 27% rated availability of appropriate ancillary staff as ‘seldom’ or ‘never’, 39% indicated availability as ‘sometimes’, and only 29% rated availability as ‘often’ or ‘always’.(28) In the context of staffing shortages and nursing burnout, organizations need to understand the optimal care team composition for both patient and worker outcomes.

### Community, hospital, and patient characteristics

Nearly half of the reviews included in the study did not address community, hospital or organizational, or patient characteristics. When hospital characteristics were included in reviews, there was little consensus on how to best define and control for characteristics that may influence the provision of high-quality care. For example, hospitals equipped to perform open heart surgeries were used as proxy for more advanced equipment and technological capacity than those who do not perform open heart surgery, but it is unclear whether this denotes a substantive difference in organizational capacity or resources compared to a hospital that does not perform open heart surgeries. Data suggests that community level factors such as urbanicity, county socioeconomic status, and housing type may shape the patient population served by the organization, which impacts both care delivery and outcomes,(10) so these factors need to be considered when examining the impacts of staffing and workforce issues.

None of the included reviews addressed SDOH as patient characteristics, nor did they use the term ‘health equity’ in describing patient outcomes. This may have occurred in part because of the temporality of the reviews and the papers they sourced, but is nonetheless a gap in our understanding of relationships between staffing and patient outcomes. In general, patient characteristics such as demographics were not consistently included in staffing models, and more nuanced patient-level metrics such as insurance status or admission type were even less commonly included. Measures intended to reflect patient acuity, such as an adjustment for comorbidities or severity of illness, were similarly sparse. In the existing literature, some studies reported diagnostic codes or acuity ratings to quantify workload related to patient care, but even these measures failed to identify patients requiring extra time and personnel resources to achieve similar outcomes.(29) This implies an over-reliance on patient demographics such as age, sex, and race as proxies for SDOH and therefore limits the interpretation of results and their impact on health equity. This pattern may be related to a lack of available data in primary research studies endeavoring to quantify these relationships. In Washington State, House Bill 1272 requires hospitals to begin reporting additional patient demographic information in 2023. This data should improve future researchers’ ability to assess health equity as a critical outcome.

### Variable definition and analytic strategy

Definitions of staffing metrics used by primary studies were rarely provided in the systematic reviews, suggesting that these variables were not consistently defined in workforce research. The lack of clear definitions or consistent measures makes further analysis and application of findings difficult. For example, the most common staffing metric used in the study was nurse-to-patient ratio. This ratio does not overtly reflect patient acuity as well as measurements such as Nursing Hours Per Patient Day, and therefore hinders the comparison of staffing ratios and patient outcomes among different populations and in different acute care settings.

Along with inconsistent variable definitions, we found a lack of consensus regarding the analytic strategy or model type used in studies exploring the impacts of healthcare staffing on patient outcomes. Most included reviews did not report the methods of quantitative analysis used by primary workforce studies. Of those reporting analytic strategies, multivariate logistic regression was the most common method of analysis. Future studies in this field could explore which model types and methods best reflect the relationship between healthcare staffing and patient outcomes, particularly when examining the influence of community, hospital, or patient characteristics.

### Limitations

A review has inherent limitations, since findings generally rely on the accuracy and completeness of the review, and quality may vary across studies. For example, reviews may have poorly specified inclusion and exclusion criteria or an inadequate search process. This study relied on the information presented in the papers we reviewed, which may have missing information from primary studies or may be interpreted differently in our findings. Additionally, despite careful development of a systematic search strategy, we may have missed some relevant systematic reviews. Despite the various limitations, a key strength of this report is the sheer volume of primary studies assessed. More than 575 studies were represented in the included reviews. Examination of reference lists showed that 13 primary studies were represented in multiple reviews, with 9/13 in 2 reviews, 3/13 in 3 reviews, and 1/13 in 4 reviews.

However, assessment of each review showed that findings were not exclusively drawn from duplicated studies. Altogether, this review offers a comprehensive overview of current studies about interprofessional team composition and staffing and their impacts on patient outcomes in the hospital setting.

## Conclusions

This review revealed that contextual factors such as healthcare team composition or hospital setting were largely unexamined in current health services literature. Further research is needed to better understand how these factors impact hospital function, work environment, care quality, and staff and patient outcomes. Given the pervasive and ongoing shortages of workers in healthcare, we need to build on our understanding of nurse staffing by examining how the availability of interprofessional care team members impact clinician workload and patient outcomes, and how hospitals in different settings and those serving diverse patient populations may require different care team composition. This work can inform strategies to optimize team composition with the goal of improving patient outcomes and furthering health equity. In addition, to promote comparability across studies, future workforce research should include a comprehensive analytic approach that includes clearly defined variables representing interprofessional care teams, community factors, and staffing and patient characteristics. More comprehensive and applicable research can better inform practice and policy, improving outcomes for patients, workers, and communities.

## Abbreviations

SDOH: Social determinants of health RN: Registered nurse
PRISMA-ScR: Preferred Reporting Items for Systematic Reviews and Meta-Analysis
PICOS: Population, Intervention, Comparison, Outcome, and Study type
MeSH: Medical Subject Heading
CINAHL: Cumulated Index to Nursing and Allied Health Literature
NPR: Nurse-to-patient ratio
NHPPD: Nurse Hours Per Patient Day
NAS: Nursing Activities Score
TISS: Therapeutic Intervention Scoring System

## Declarations

### Ethics approval and consent to participate

Not applicable

### Consent for publication

Not applicable

### Availability of data and materials

The datasets used and/or analyzed during the current study are available from the corresponding author on reasonable request.

### Authors’ contributions

AP is the corresponding author for this study. AP and ZG designed and performed the literature extraction. CM executed the literature search. AP and KW drafted the manuscript and designed the figures. NBS, NH, PL, AF, and SI were involved in planning and supervising the work. All authors reviewed and contributed to editing the manuscript.

### Data Availability

Data used in this review are available in Appendix 1 (Supporting Information 1).

## Acknowledgments

Not applicable.

## References

1. American Nurses Foundation. Pulse on the nation’s nurses COVID-19 series. American Nurses Foundation.; 2020. https://www.nursingworld.org/practice-policy/work-environment/health-safety/disaster-preparedness/coronavirus/what-you-need-to-know/mental-health-and-wellbeing-survey/. Accessed 20 Mar 2022.

2. Brown KM, Robinson GE, Nadelson CC, Grigoriadis S, Mittal LP, Conteh N, et al. Psychological impact of COVID-19 on minority women. J Nerv Mental Dis. 2021;209(10):695–6.

3. Zipf AL, Polifroni EC, Beck CT. The experience of the nurse during the COVID-19 pandemic: A global meta-synthesis in the year of the nurse. J Nurs Scholarship. 2022;54(1):92–103.

4. Boone LD, Rodgers MM, Baur A, Vitek E, Epstein C. An integrative review of factors and interventions affecting the well-being and safety of nurses during a global pandemic. Worldv Evid-Based Nu. 2023;20:107–115.

5. Riman KA, Harrison JM, Sloan DM, McHugh MD. Work Environment and Operational Failures Associated With Nurse Outcomes, Patient Safety, and Patient Satisfaction. Nursing Research. 2023;72(1):p 20–29.

6. Dierkes A, Do D, Morin H, Rochman M, Sloane DM, McHugh MD. The impact of California’s staffing mandate and the economic recession on registered nurse staffing levels: A longitudinal analysis. Nurs Outlook. 2022;70(2):219–27.

7. Pittman P. Evidence on hospital staffing and outcomes: implications for Washington. Washington, DC: George Washington University; 2022.

8. Griffiths P, Recio-Saucedo A, Dall’Ora C, Briggs J, Maruotti A, Meredith P, et al. The association between nurse staffing and omissions in nursing care: a systematic review. J Adv Nurs. 2018;74(7):1474–87.

9. Gottlieb LN, Gottlieb B, Bitzas V. Creating empowering conditions for nurses with workplace autonomy and agency: how healthcare leaders could be guided by strengths-based nursing and healthcare leadership (SBNH-L). J Healthcare Leadership. 2021;13:169.

10. Al-Amin M, Islam MN, Li K, Shiels N, Buresh J. Is there an association between hospital staffing levels and inpatient-COVID-19 mortality rates? PLoS One. 2022;17(10):e0275500.

11. Wammes JJG, van der Wees PJ, Tanke MAC, Westert GP, Jeurissen PPT. Systematic review of high-cost patients’ characteristics and healthcare utilisation. BMJ Open. 2018;8(9):e023113.

12. Washington state legislature. E2SHB 1272 bill report: Concerning health system transparency. https://lawfilesext.leg.wa.gov/biennium/2021-22/Pdf/Bill%20Reports/House/1272-S2.E%20HBR%20FBR%2021.pdf?q=20231219104654. Accessed 19 Dec 2023.

13. Armstrong R, Hall BJ, Doyle J, Waters E. ‘Scoping the scope’of a cochrane review. J Pub Hlth. 2011;33(1):147–50.

14. Levac D, Colquhoun H, O’Brien KK. Scoping studies: advancing the methodology. Impl Sci. 2010;5:1–9.

15. Page MJ, McKenzie JE, Bossuyt PM, Boutron I, Hoffmann TC, Mulrow CD, et al. The PRISMA 2020 statement: an updated guideline for reporting systematic reviews. Sys Rev. 2021;10(1):89.

16. Bagnasco A, Dasso N, Rossi S, Galanti C, Varone G, Catania G, et al. Unmet nursing care needs on medical and surgical wards: A scoping review of patients’ perspectives. J Clin Nurs. 2020;29(3-4):347–69.

17. Dall’Ora C, Saville C, Rubbo B, Turner L, Jones J, Griffiths P. Nurse staffing levels and patient outcomes: A systematic review of longitudinal studies. Int J Nurs Stud. 2022:104311.

18. Rae PJ, Pearce S, Greaves PJ, Dall’Ora C, Griffiths P, Endacott R. Outcomes sensitive to critical care nurse staffing levels: A systematic review. Intensive Crit Care Nurs. 2021;67:103110.

19. Plotnikoff KM, Krewulak KD, Hernández L, Spence K, Foster N, Longmore S, et al. Patient discharge from intensive care: an updated scoping review to identify tools and practices to inform high-quality care. Crit Care. 2021;25(1):1–13.

20. Cassarino M, Robinson K, Quinn R, Naddy B, O’Regan A, Ryan D, et al. Impact of early assessment and intervention by teams involving health and social care professionals in the emergency department: A systematic review. PLoS One. 2019;14(7):e0220709.

21. Kerlin MP, Adhikari NK, Rose L, Wilcox ME, Bellamy CJ, Costa DK, et al. An official American Thoracic Society systematic review: the effect of nighttime intensivist staffing on mortality and length of stay among intensive care unit patients. Am J Resp Crit Care Med. 2017;195(3):383–93.

22. Evangelou E, Lambrinou E, Kouta C, Middleton N. Identifying validated nursing quality indicators for the intensive care unit: an integrative review. Connect: The World of Crit Care Nurs. 2018;12(2):28–39.

23. Burke JR, Downey C, Almoudaris AM. Failure to rescue deteriorating patients: a systematic review of root causes and improvement strategies. J Patient Safety. 2022;18(1):e140–e55.

24. McGahan M, Kucharski G, Coyer F. Nurse staffing levels and the incidence of mortality and morbidity in the adult intensive care unit: a literature review. Australian Crit Care. 2012;25(2):64–77.

25. Recio-Saucedo A, Dall’Ora C, Maruotti A, Ball J, Briggs J, Meredith P, et al. What impact does nursing care left undone have on patient outcomes? Review of the literature. J Clin Nurs. 2018;27(11-12):2248–59.

26. Bae S-H, Fabry D. Assessing the relationships between nurse work hours/overtime and nurse and patient outcomes: systematic literature review. Nurs Outlook. 2014;62(2):138–56.

27. National Quality Forum. Serious reportable events in healthcare-2011 update: a concensus report. Washington, DC: National Quality Forum; 2011.

28. American Nurses Foundation. COVID-19 survey series: 2022 workplace survey. American Nurses Foundation; 2022. https://www.nursingworld.org/practice-policy/work-environment/health-safety/disaster-preparedness/coronavirus/what-you-need-to-know/covid-19-survey-series-anf-2022-workplace-survey/. Accessed 1 Nov 2022.

29. Juvé-Udina M-E, Adamuz J, López-Jimenez M-M, Tapia-Pérez M, Fabrellas N, Matud-Calvo C, et al. Predicting patient acuity according to their main problem. J Nurs Manage. 2019;27(8):1845–58.

30. Miranda DR, de Rijk A, Schaufeli W. Simplified Therapeutic Intervention Scoring System: the TISS-28 items--results from a multicenter study. Crit Care Med. 1996;24(1):64–73.

31. Miranda DR, Nap R, de Rijk A, Schaufeli W, Iapichino G. Nursing Activities Score. Crit Care Med. 2003;31(2):374–82.

